# Temporal change in etiology and clinical characteristics of hepatocellular carcinoma in a large cohort of patients with hepatocellular carcinoma in New South Wales, Australia

**DOI:** 10.1101/2023.02.20.23286164

**Authors:** Yuen Kang Joseph Yeoh, Gregory J. Dore, Ian Lockart, Mark Danta, Ciara Flynn, Conner Blackmore, Miriam T Levy, Jacob George, Maryam Alavi, Behzad Hajarizadeh

## Abstract

**Background:** Viral hepatitis and alcohol-related liver disease (ALRD) are the main risk factors for hepatocellular carcinoma (HCC) in many countries. In Australia, given the access to hepatitis C virus (HCV) direct-acting antiviral (DAA) therapy since 2016, a temporal change in HCC etiology was hypothesized. This study evaluated the temporal change in the etiology and characteristics of HCC in New South Wales (NSW).

**Methods:** Patients diagnosed with HCC, admitted to three public hospitals in NSW between 2008-21, were included in analyses. We assessed the annual frequency of each HCC etiology and the distribution of HCC characteristics in participants.

**Results:** Among 1,370 patients, the most common HCC etiologies were HCV (n=483, 35%), ALRD (n=452, 33%), non-alcoholic fatty liver disease (n=347, 25%), and hepatitis B virus (n=301, 22%). The proportion of HCV-related HCC was the highest in 2011-16 (41%), and significantly declined to 30% in 2017-21 (OR: 0.53, 95%CI 0.35–0.79; p=0.002). The proportion of HCC with earlier diagnosis (BCLC stage O/A) increased from 41% in 2008-09 to 56% in 2020-21 (OR per annum: 1.05; 95%CI: 1.02–1.08; p=0.002), and proportion of patients receiving curative HCC management increased from 29% in 2008-09 to 41% in 2020-21 (OR per annum: 1.06; 95%CI: 1.03–1.10; p<0.001).

**Conclusion:** The contribution of HCV to HCC burden has been decreasing in the DAA era, suggesting the role of HCV elimination in decreasing HCC risk. Increasing frequency of less advanced HCC at diagnosis over time suggests improved HCC surveillance.

**Lay Summary:** In New South Wales, the trend of liver cancer caused by hepatitis C increased during 2008-2016, and then significantly declined after 2016, most probably due to wide access to new hepatitis C treatment (direct-acting antiviral therapy). During 2008-2021, the proportion of patients with liver cancer whose cancer was diagnosed at early stage and proportion of those receiving curative liver cancer management increased.

## Introduction

In 2020, primary liver cancer was the second leading cause of cancer death globally with more than 800,000 deaths,^1^ with its incidence projected to increase by 2030.^2^ Hepatocellular carcinoma (HCC), is the most common type of primary liver cancer.^1,3^ In Australia, while the mortality of most cancers has been decreasing in the past 20 years, the age-standardized mortality rate of HCC has almost doubled, increasing from 4.0 to 7.4 per 100,000 between 2001 and 2021.^4^

The main risk factors of HCC in Australia include hepatitis C virus (HCV), hepatitis B virus (HBV), alcohol-related liver disease (ARLD) and metabolic-related liver disease.^5-7^ Direct-acting antiviral therapies (DAA) are highly curative treatments for HCV.^8,9^ Previous studies demonstrated a decreased risk of HCC following DAA therapy among people with HCV.^10,11^ HBV antiviral therapies have also decreased the risk of HCC among people with HBV.^12,13^ Given the availability of entecavir and tenofovir, as highly potent HBV antiviral therapies, in Australia since 2006,^5^ and HCV DAA therapy since 2016,^14^ a temporal change in the etiology of HCC is hypothesized. However, limited data are available to evaluate this hypothesis. Furthermore, international and Australian guidelines recommend HCC surveillance among people at greater risk of HCC,^15-17^ but the data evaluating the implementation of these recommendations and the impact on early diagnosis of HCC at population-level are limited. This study was conducted to evaluate the temporal change in etiology, and characteristics of HCC among people diagnosed with HCC in three large public hospitals in New South Wales (NSW), Australia.

## Methodology

### Study design and population

This was a retrospective cohort study of patients diagnosed with incident HCC between January 2008 and December 2021 and managed in three large public hospitals in NSW, Australia, including St Vincent’s, Liverpool, and Westmead hospitals.

All patients with the diagnostic criteria of HCC based on the 10^th^ revision of the International Classification of Diseases (ICD-10) code for liver cell carcinoma (C22.0)^18^ were identified through existing hospital databases. Patients who had a confirmed HCC diagnosis within the study period and had received their initial HCC management at one of the three hospitals involved were included. Patients with a diagnosis of combined HCC-cholangiocarcinoma or fibrolamellar-HCC were excluded.

The study was approved by the St Vincent’s Hospital, Sydney Human Research Ethics Committee (2019/STE16231), and the local ethics committees of the involved hospital sites. The approval included patient consent waivers and met the ethical guidelines of the Declaration of Helsinki. The study involved the use of the UNSW Research Electronic Data Capture, as a secure online database, to ensure patient confidentiality.

### Study procedures

A database had been developed including data of eligible patients, diagnosed between 2008 and 2019.^19,20^ In this study, this database was updated to include data to December 2021. Data collection was retrospective and involved reviewing multidisciplinary team reports, clinical notes, medication lists, and results of laboratory, histopathology, and medical imaging assessments. Collected variables included patients’ demographic and clinical characteristics. Demographic variables included age, gender, country of birth, and Australian Indigenous ethnicity. Clinical variables included HCC diagnosis date, cause of background liver disease, treatment of background liver disease, diabetes, Child-Pugh class, Model for End-stage Liver Disease (MELD) score, decompensated cirrhosis (identified as presentation of ascites, hepatic encephalopathy, or portal hypertensive bleeding), alpha-fetoprotein (AFP) level, Barcelona Clinic Liver Cancer (BCLC) tumor stage, size and number of lesions, Milan criteria, surveillance detection, initial HCC management, and survival related data.

### Study definitions

Patients were classified based on their HCC etiologies, ascertained from records in the clinical notes and laboratory results. Patients were classified as HCV-related HCC (HCV HCC) if they had a record of HCV diagnosis, confirmed by a positive HCV RNA, or had previously received HCV therapy. Patients were classified as HBV-related HCC (HBV HCC) if they had been diagnosed with a positive HBV surface antigen or had previously received HBV therapy. Etiologies including ARLD, non-alcoholic fatty liver disease (NAFLD) and non-alcoholic steatohepatitis (NASH) were identified based on multidisciplinary team reports. Although the current terminology of NAFLD includes NASH as a sub-group,^21^ we used NAFLD/NASH as a compound phrase for HCC etiology given various terminology used in clinical notes, particularly in the early years of this study. Other etiologies like autoimmune hepatitis, primary biliary cholangitis, primary sclerosing cholangitis, haemochromatosis, alpha-1-antitrypsin deficiency, or other liver diseases, were categorized as ‘Others’ due to relatively small numbers. Patients with no documented underlying liver diseases were categorized as ‘Nil/Unknown’ etiology.

For patients with more than one etiology, different strategies were used for descriptive analysis and trend analysis. For descriptive analysis, multiple HCC etiologies were allowed, with each patient included in each relevant etiology category (Table 1). For trend analysis, patients were grouped into a single etiology category to be able to assess the mutually exclusive proportion of each etiology in each year. For trend analyses, HCV HCC included patients with HCV, with or without any other etiology (patients with HCV and co-existing ARLD were further sub-grouped as HCV-ARLD). HBV HCC included patients with HBV, with or without any other etiology (except for HCV, as described above). Patients with ARLD and NAFLD/NASH were grouped under ARLD, as ARLD was most likely to be the primary etiology. NAFLD/NASH HCC included patients with NAFLD/NASH and no other etiologies mentioned above. Patients who were not categorized in any of the above-mentioned categories were grouped as ‘Others’.

**Table 1:**
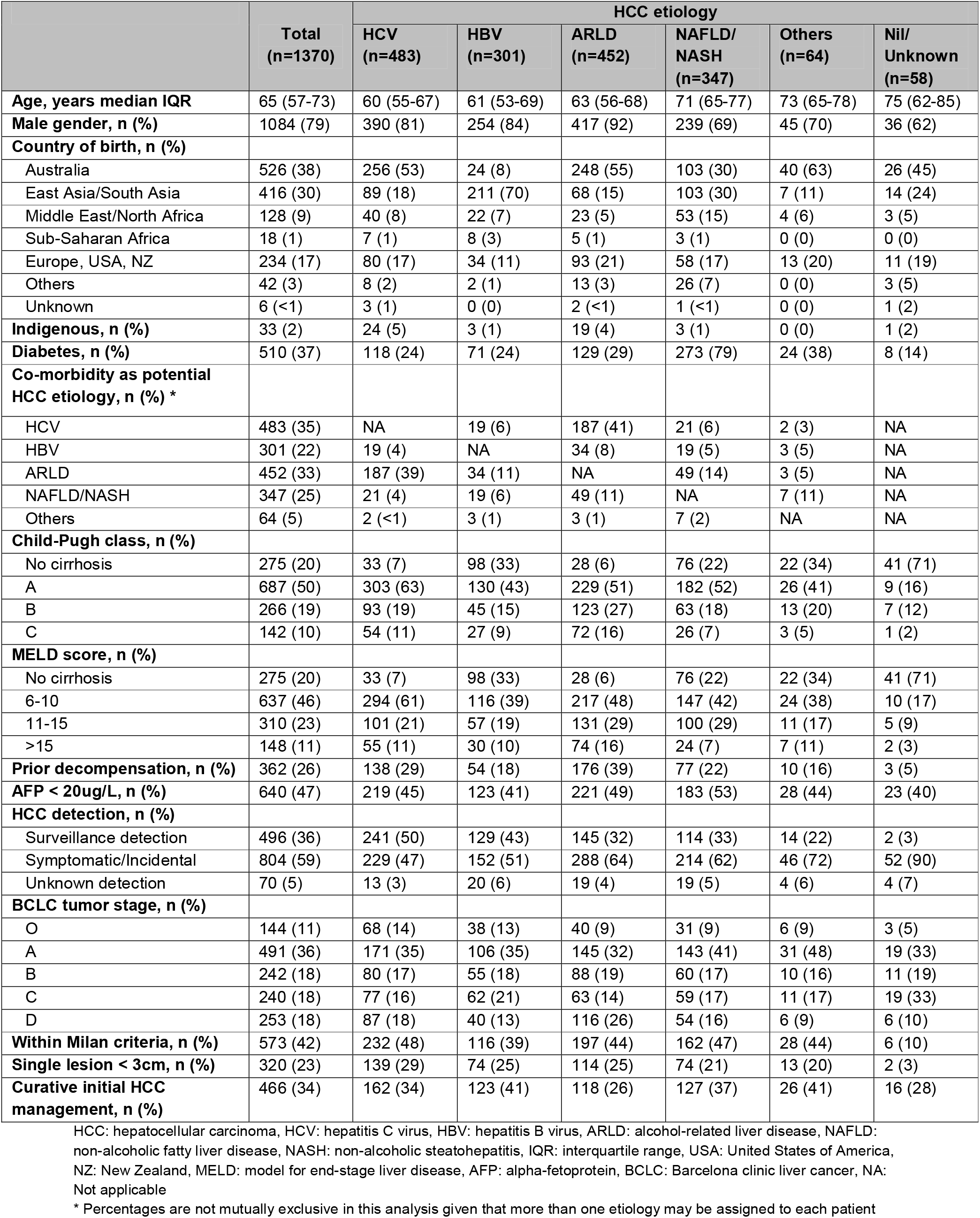
Baseline characteristics of patients with HCC, overall and by HCC etiology

The date of HCC diagnosis was defined as the date of identification of HCC by multiphase imaging or the date of biopsy if imaging was not used. Age was calculated from the date of birth to the date of HCC diagnosis. Cirrhosis was categorized based on Child-Pugh class and MELD score, obtained from clinical, pathology and blood test results. History of decompensation (i.e., hepatic encephalopathy, ascites or portal hypertensive bleeding) was extracted from either imaging reports or medical records. Surveillance HCC detection was defined as incident HCC in patients who were asymptomatic and underwent imaging for the purpose of HCC screening. Patients were considered as managed by curative treatment methods if they received a liver transplant, surgical liver resection, or liver ablation.

### Statistical analysis

Distribution of demographic and clinical characteristics was evaluated by HCC etiology and year of HCC diagnosis. Given that HCV DAA treatment has been government-funded and broadly available since March 2016 in Australia,^14^ an interrupted time series regression model^22^ was used to evaluate changes in the trend of the proportion of HCV HCC in each year during 2008-16 compared with 2017-21. Generalized linear models were used to evaluate the trend of the annual distribution of demographic and clinical characteristics of participants, assuming a binomial distribution of the dependent variables and a logit transformation as the link function. Demographic characteristics, used as dependent variables, included proportions of men, men older than 65 years and women older than 65 years. Clinical characteristics, used as dependent variables, included proportions of Child-Pugh class B/C, MELD score>11, decompensated cirrhosis, HCC detected by surveillance, HCC BCLC stage O/A, and curative initial HCC management. A p-value <0.05 was considered statistically significant. All analyses were performed using STATA (v16.1; Stata Corp, College Station, TX).

## Results

### Baseline characteristics

From 2008 to 2021, a total of 1,394 patients were diagnosed with HCC and managed in the three hospitals. Of these, three patients with insufficient medical records, 16 with combined HCC-cholangiocarcinoma and five with fibrolamellar-HCC were excluded. The remaining 1,370 patients were included in the analysis, with baseline characteristics summarized in Table 1. The median age was 65 years, and 79% were men. The most common HCC etiologies were HCV (n=483, 35%), ARLD (n=452, 33%), NAFLD/NASH (n=347, 25%), and HBV (n=301, 22%). In 36% of patients, HCC was detected through surveillance. At the time of HCC diagnosis, 20% of patients did not have cirrhosis, 26% had experienced at least one episode of decompensation, and 47% were in the BCLC stage of O or A.

Among patients with HBV HCC, 70% were born in East or South Asia, and 33% did not have cirrhosis at the time of HCC diagnosis. Among patients with NAFLD/NASH HCC, the median age was 71 and 79% had diabetes, while 22% did not present with cirrhosis at the time of HCC diagnosis.

Among patients with HCV HCC, 39% had ARLD as further potential HCC etiology. Among patients with ARLD HCC, 41% had HCV, and 11% had NAFLD/NASH as further potential HCC etiologies.

Among patients with HCV and HBV HCC, 50% and 43%, respectively, were diagnosed through screening, higher than 32% among patients with ARLD and 33% among patients with NAFLD/NASH.

### Annual trends in HCC etiology

The annual number of patients with HCC increased from 53 in 2008 to 146 in 2019, while the number dropped sharply after 2019 to 106 in 2020 and 73 in 2021 (Figure 1A).

**Figure 1:**
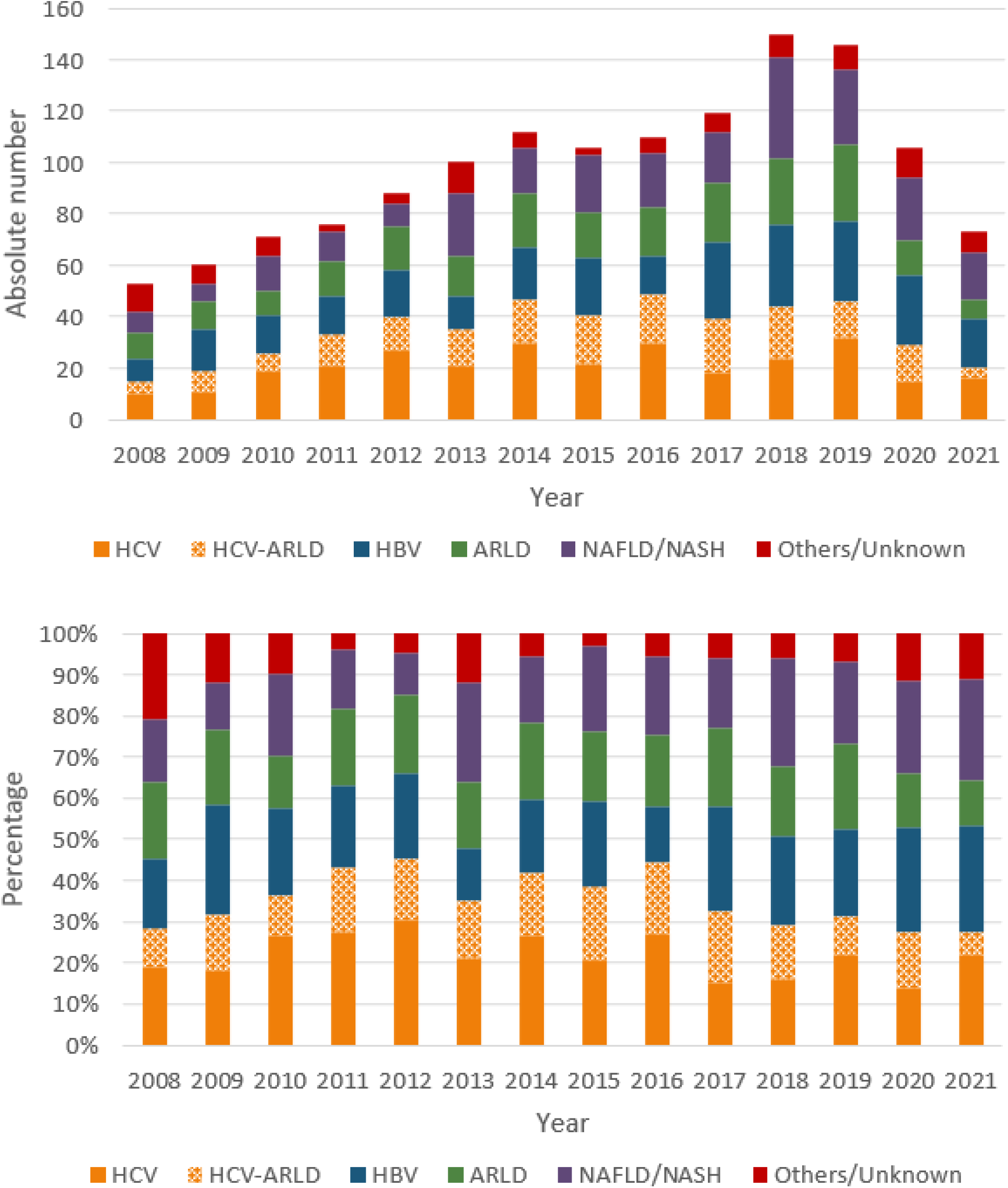
Absolute number (A) and proportion (B) of patients with HCC in each year during 2008-21, by HCC etiology. (HCC: hepatocellular carcinoma, HCV: hepatitis C virus, ARLD: alcohol-related liver disease, HBV: hepatitis B virus, NAFLD: non-alcoholic fatty liver disease, NASH: non-alcoholic steatohepatitis)

The proportion of patients with HCV (i.e., HCV HCC and HCV-ARLD HCC) increased steadily from 28% in 2008 to 44% in 2012, remained relatively stable until 2016 (44%), then continuously decreased afterward to 34% in 2017 and 25% in 2021 (Figure 1B). The interrupted time series analysis demonstrated a significant change in the trend of HCV HCC after 2016 [odds ratio (OR): 0.53; 95% CI 0.35–0.79; p=0.002; Figure 2]. Moreover, the number of patients with HCV HCC achieving sustained virologic response due to HCV treatment has increased since 2016 (Supplementary Figure 1).

**Figure 2:**
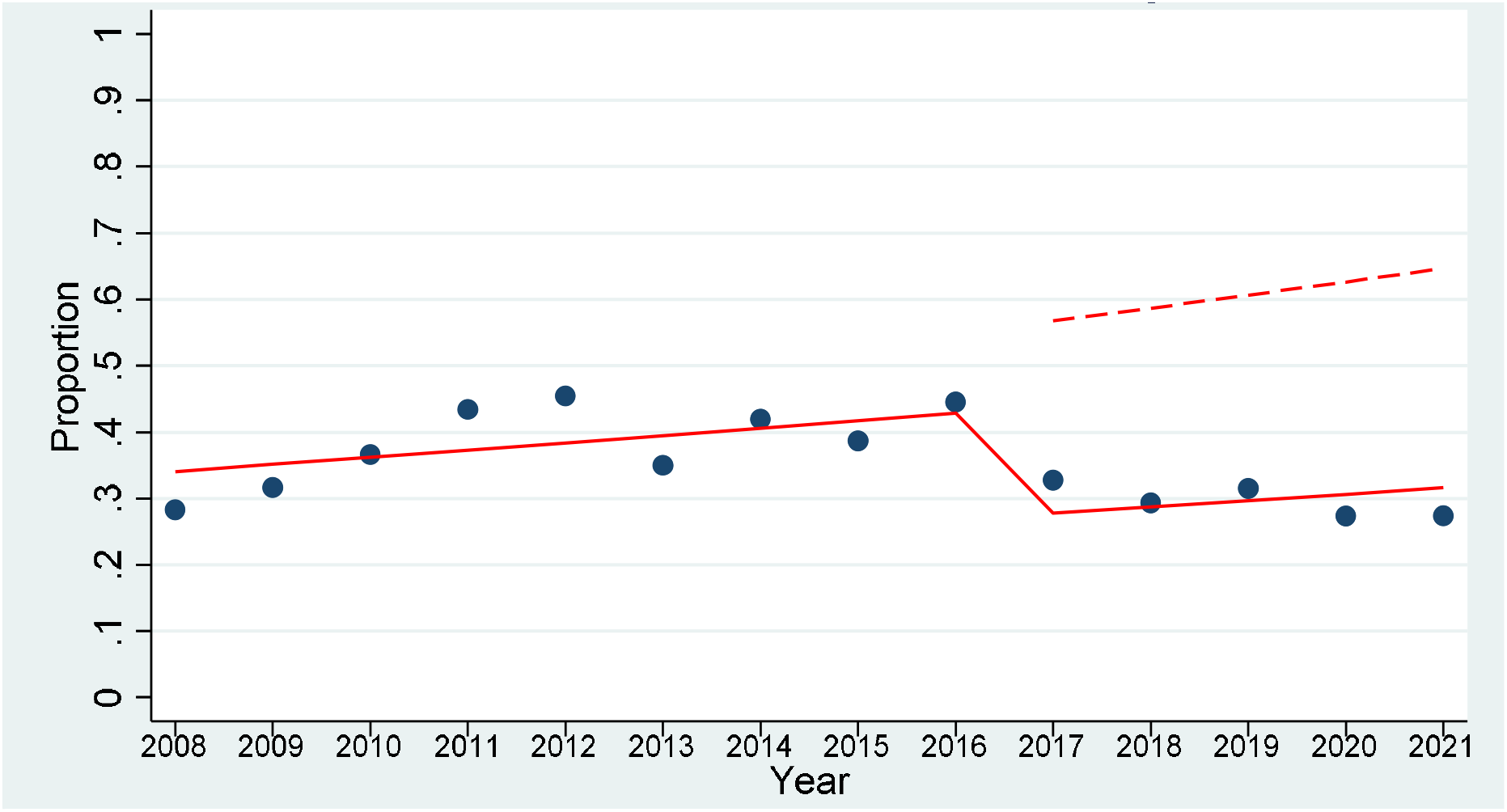
Proportion of HCV-related HCC cases during 2008-21. (The solid red line represents the actual trend over time and the dashed red line represents the predicted trend, assuming the pre-2016 trend continued) (HCC: hepatocellular carcinoma, HCV: hepatitis C virus)

The distribution of HCC etiology over time are shown in Table 2 and Figure 1. The trend analyses for HCC etiologies other than HCV included the period between 2008 and 2016, given that the significant decrease in HCV HCC post-2016 affected the proportion of all other HCC etiologies. Among other etiologies, NAFLD/NASH showed an increasing trend with the proportion of NAFLD/NASH HCC increasing from 13% during 2008-9 to 19% in 2016 although the trend was not statistically significant (OR per annum: 1.05; 95% CI 0.97–1.14; p=0.200). The proportion of HBV HCC decreased from 22% in 2008-09 to 14% in 2016 although the trend was not statistically significant (OR per annum: 0.95; 95% CI 0.88–1.02; p=0.166). The proportion of ARLD HCC remained relatively stable during 2008-16 (OR per annum: 1.04; 95% CI 0.98–1.11; p=0.194).

**Table 2:** Baseline characteristics of patients with HCC, by year of HCC diagnosis

|  | 2008-09<br>(n=113) | 2010-11<br>(n=147) | 2012-13<br>(n=188) | 2014-15<br>(n=218) | 2016-17<br>(n=229) | 2018-19<br>(n=296) | 2020-21<br>(n=179) |
| --- | --- | --- | --- | --- | --- | --- | --- |
| **Age, years median IQR** | 65 (57-74) | 63 (55-72) | 63 (56-72) | 64 (57-72) | 65 (58-73) | 66 (59-73) | 67 (60-76) |
| **Male gender, n (%)** | 91 (81) | 117 (80) | 152 (81) | 179 (82) | 182 (79) | 232 (78) | 131 (73) |
| **Indigenous, n (%)** | 0 (0) | 3 (2) | 1 (1) | 8 (4) | 7 (3) | 7 (2) | 7 (4) |
| **Diabetes, n (%)** | 37 (33) | 51 (35) | 63 (34) | 82 (38) | 91 (40) | 113 (38) | 73 (41) |
| **HCC etiology, n (%)** |  |  |  |  |  |  |  |
| HCV | 21 (19) | 40 (27) | 48 (26) | 52 (24) | 48 (21) | 56 (19) | 31 (17) |
| HCV-ARLD | 13 (12) | 19 (13) | 27 (14) | 36 (17) | 40 (17) | 34 (11) | 18 (10) |
| HBV | 25 (22) | 30 (20) | 31 (16) | 42 (19) | 45 (20) | 63 (21) | 46 (26) |
| ARLD | 21 (19) | 23 (16) | 33 (18) | 39 (18) | 42 (18) | 56 (19) | 22 (12) |
| NAFLD/NASH | 15 (13) | 25 (17) | 33 (18) | 40 (18) | 41 (18) | 68 (23) | 42 (23) |
| Others/Unknown | 18 (16) | 10 (7) | 16 (9) | 9 (4) | 13 (6) | 19 (6) | 20 (11) |
| **Child-Pugh class, n (%)** |  |  |  |  |  |  |  |
| No cirrhosis | 23 (20) | 31 (21) | 44 (23) | 42 (19) | 38 (17) | 62 (21) | 33 (18) |
| A | 61 (54) | 72 (49) | 90 (48) | 99 (45) | 126 (55) | 140 (47) | 100 (56) |
| B | 21 (19) | 22 (15) | 35 (19) | 47 (22) | 49 (21) | 56 (19) | 37 (21) |
| C | 8 (7) | 22 (15) | 19 (10) | 30 (14) | 16 (7) | 38 (13) | 9 (5) |
| **MELD score, n (%)** |  |  |  |  |  |  |  |
| No cirrhosis | 23 (20) | 31 (21) | 44 (23) | 42 (19) | 38 (17) | 62 (21) | 33 (18) |
| 6-10 | 55 (49) | 63 (43) | 85 (45) | 97 (44) | 125 (55) | 134 (45) | 80 (45) |
| 11-15 | 25 (22) | 40 (27) | 33 (18) | 54 (25) | 49 (21) | 64 (22) | 45 (25) |
| >15 | 10 (9) | 13 (9) | 26 (14) | 25 (11) | 17 (7) | 36 (12) | 21 (12) |
| **Prior decompensation, n (%)** | 30 (34) | 43 (37) | 51 (35) | 66 (38) | 61 (32) | 86 (37) | 25 (18) |
| **AFP < 20ug/L, n (%)** | 47 (42) | 61 (38) | 64 (39) | 106 (49) | 110 (49) | 157 (53) | 87 (49) |
| **Surveillance detection, n (%)** | 32 (28) | 51 (34) | 60 (32) | 75 (34) | 100 (44) | 102 (34) | 76 (42) |
| **BCLC tumor stage, n (%)** |  |  |  |  |  |  |  |
| O | 12 (11) | 11 (8) | 19 (10) | 19 (9) | 29 (13) | 37 (13) | 17 (10) |
| A | 34 (30) | 53 (36) | 52 (28) | 87 (40) | 77 (34) | 105 (35) | 83 (46) |
| B | 23 (20) | 25 (17) | 40 (21) | 30 (14) | 41 (18) | 35 (12) | 48 (27) |
| C | 20 (18) | 25 (17) | 38 (20) | 32 (15) | 42 (18) | 57 (19) | 26 (15) |
| D | 24 (21) | 33 (22) | 39 (21) | 50 (23) | 40 (17) | 62 (21) | 5 (3) |
| **Within Milan criteria, n (%)** | 42 (37) | 57 (39) | 65 (35) | 96 (44) | 100 (44) | 131 (44) | 82 (46) |
| **Single lesion < 3cm, n (%)** | 23 (20) | 24 (16) | 33 (18) | 61 (28) | 62 (27) | 79 (27) | 38 (21) |
| **Curative initial HCC management, n (%)** | 33 (29) | 39 (27) | 45 (24) | 76 (35) | 90 (39) | 110 (37) | 73 (41) |

HCC: hepatocellular carcinoma, IQR: interquartile range, HCV: hepatitis C virus, ARLD: alcohol-related liver disease, HBV: hepatitis B virus, NAFLD: non-alcoholic fatty liver disease, NASH: non-alcoholic steatohepatitis, MELD: model for end-stage liver disease, AFP: alpha-fetoprotein, BCLC: Barcelona clinic liver cancer

Among total patients with HCV HCC, 52% (n=253) received HCV treatment before HCC diagnosis, and 25% (n=120) had sustained virologic response (SVR) at the time of HCC diagnosis (Supplementary Figure 2). The proportion of stage O/A HCC cases at the time of diagnosis was 66% and 31% among participants receiving HCV treatment and those not receiving treatment, respectively. Among total patients with HBV HCC, 48% (n=135) were receiving HBV treatment prior to HCC diagnosis, with the annual proportion fluctuating (Supplementary Figure 2). The proportion of HBV HCC cases with BCLC stage O/A at the time of diagnosis was 68% and 27% among participants receiving HBV treatment and those not receiving treatment, respectively.

### Trends in age and gender distribution

The proportion of men was relatively stable between 2008 and 2018 (OR per annum: 0.97; 95% CI 0.93–1.00; p=0.068; Table 2, Figure 3). The proportion of men older than 65 years increased from 48% in 2008 to 59% in 2021 (OR per annum: 1.04; 95% CI 1.01–1.08; p=0.016; Figure 3). No significant trend was found in the proportion of women older than 65 years (OR per annum: 0.99; 95% CI 0.93–1.06; p=0.760; Figure 3).

**Figure 3:**
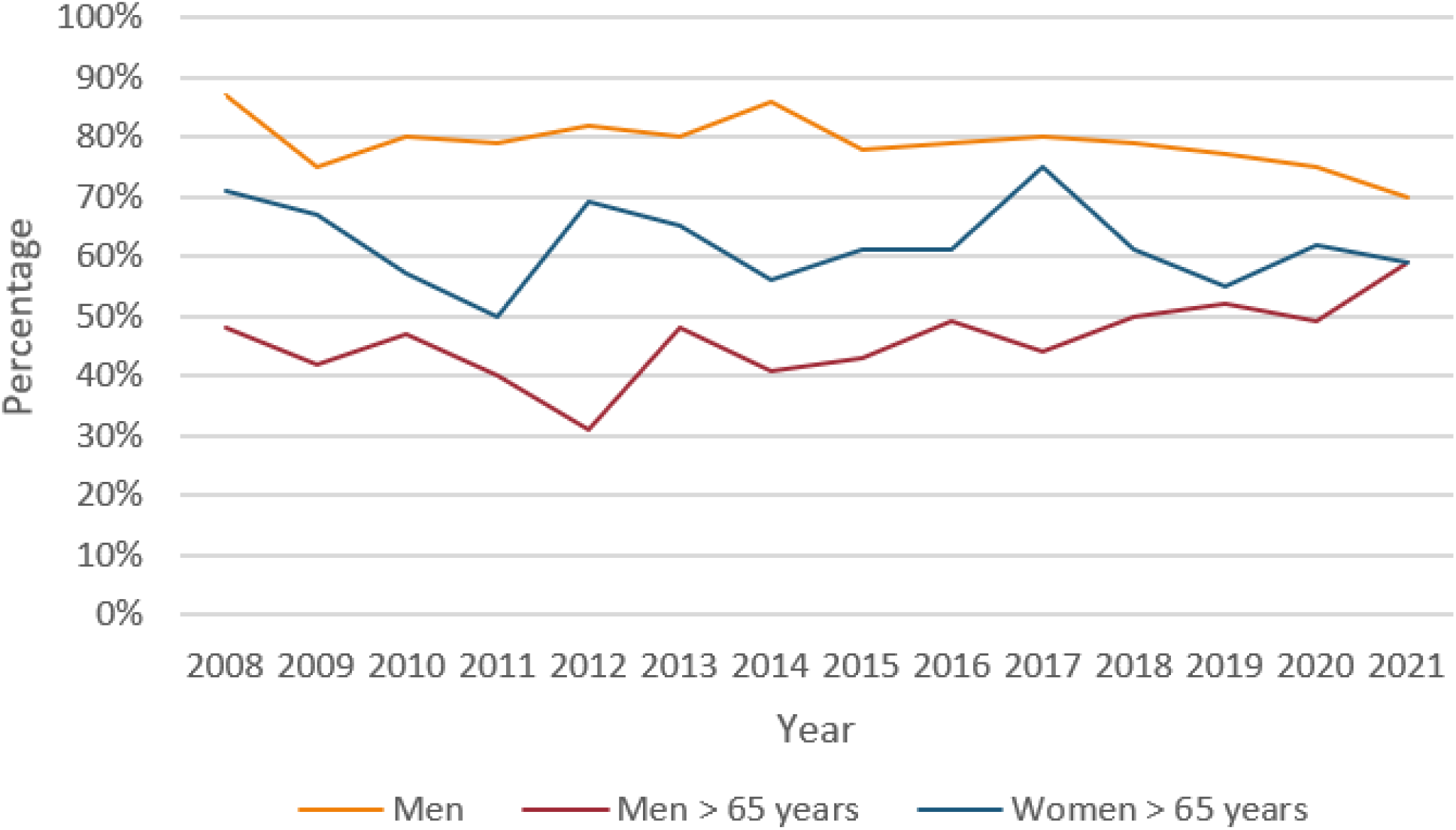
Proportion of patients with HCC in each year during 2008-21, by gender and age.

### Trends in underlying liver disease and HCC characteristics

The distribution of underlying liver disease in participants over time is shown in Table 2 and Figure 4. A decreasing trend over time was identified in the proportion of patients with prior decompensation (OR per annum: 0.96; 95% CI: 0.93–0.99; p=0.027) although this result should be interpreted cautiously given the fluctuating pattern (Figure 4). No significant trend was found in the proportion of patients with Child-Pugh class B/C (OR per annum: 1.00; 95% CI: 0.97–1.04); p=0.793) and the proportion of patients with MELD score>11 (OR per annum: 0.99; 95% CI: 0.96–1.02; p=0.457).

**Figure 4:**
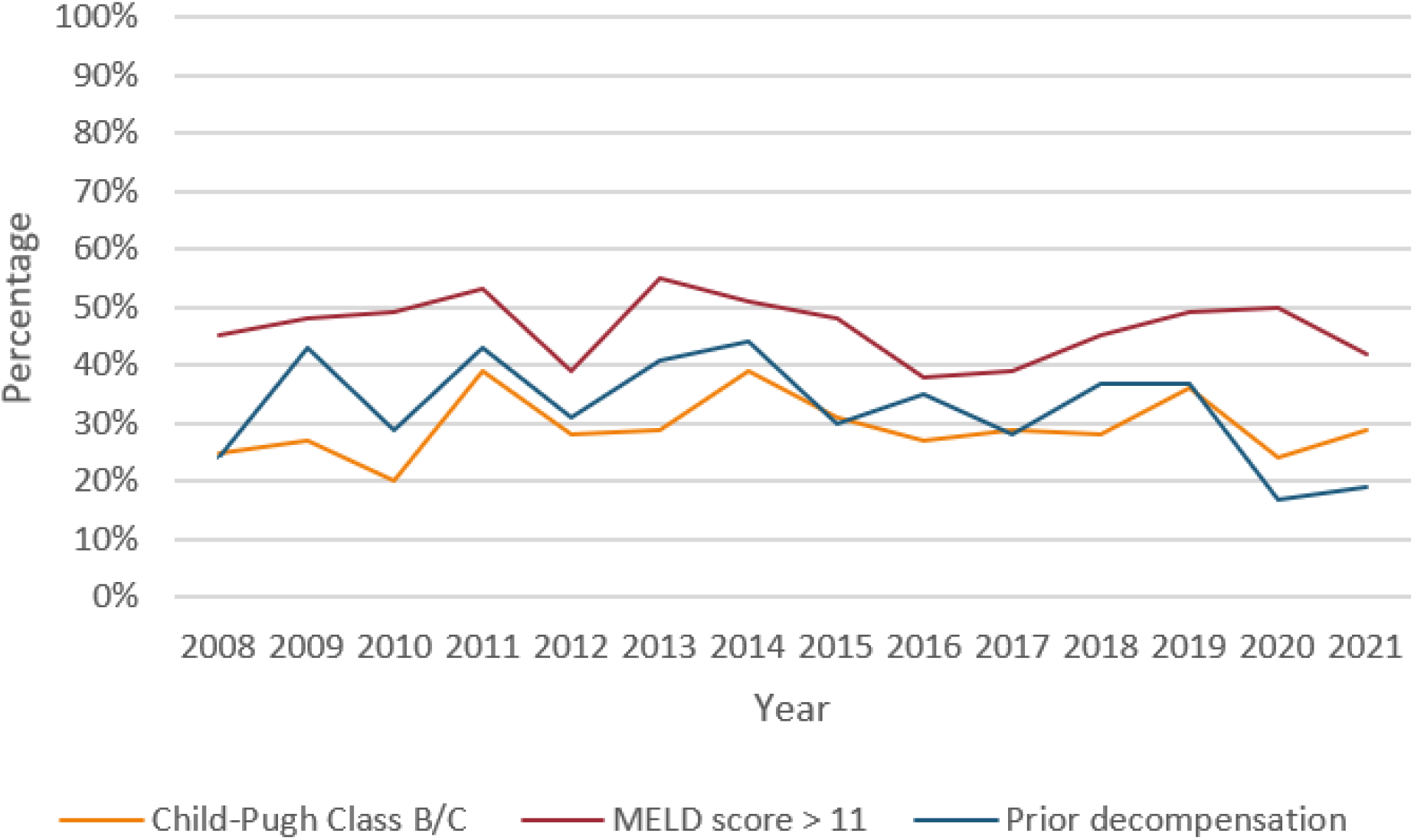
Proportion of patients with HCC in each year during 2008-21, by underlying liver disease severity. (MELD: model for end-stage liver disease)

The distribution of HCC characteristics in participants over time was shown in Table 2 and Figure 5. The proportion of HCC cases detected through surveillance increased from 28% in 2008-9 to in 44% in 2016-17, temporarily decreased in 2018-19, and then increased again to 42% in 2020-21 (OR per annum: 1.02; 95% CI: 0.99–1.05; p=0.186). However, more HCC cases were diagnosed at an earlier stage, as evidenced by a significant increase in the proportion with BCLC stage O/A from 41% in 2008-09 to 56% in 2020-21 (OR per annum: 1.05; 95% CI: 1.02–1.08; p=0.002). The patients were also more likely to receive initial HCC curative management in recent years, with 29% of patients in 2008-09 compared to 41% in 2020-21 receiving initial HCC curative management (OR per annum: 1.06; 95% CI: 1.03–1.10; p<0.001).

**Figure 5:**
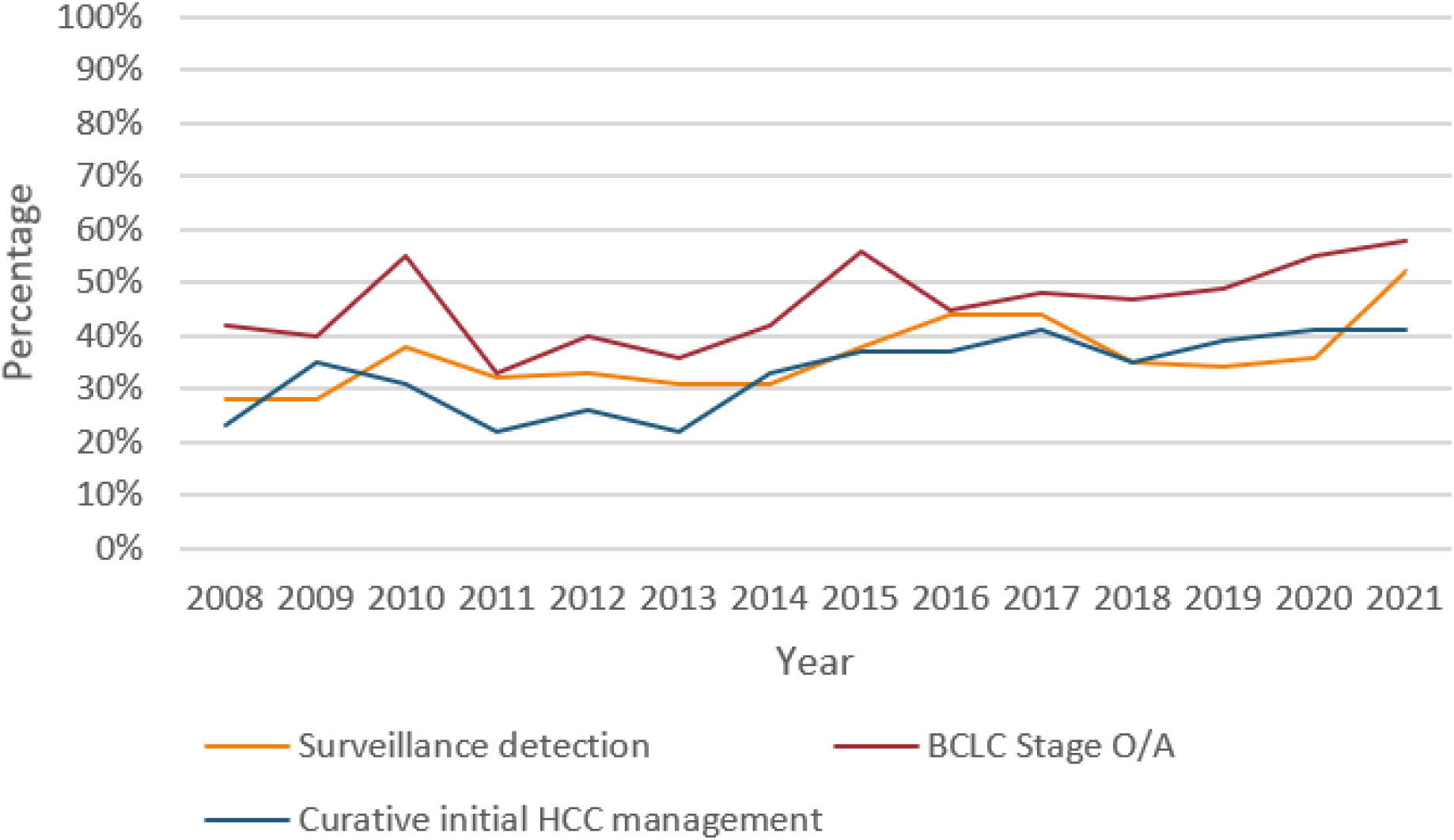
Proportion of patients with HCC in each year during 2008-21, by clinical characteristics of HCC. (BCLC: Barcelona clinic liver cancer, HCC: hepatocellular carcinoma)

## Discussion

This study demonstrated that the overall number of HCC cases decreased after 2019. The proportion of patients with HCV HCC decreased after 2016, following broad access to DAA therapy. There was also a temporal trend to earlier HCC diagnosis, with more patients eligible for initial curative management. These data improve our understanding of the trends in HCC etiology and characteristics in NSW, which could inform healthcare policies to improve the prevention and management of HCC.

Our data demonstrated a steady increase in the annual number of HCC cases from 2008 to 2019, followed by a sharp decline after 2019. The initial increasing trend is consistent with the data projecting an increasing HCC incidence in Australia through 2030.^2^ There are two potential hypotheses for declining HCC cases after 2019. First, with the COVID-19 pandemic overwhelming healthcare systems and other restrictions, many clinical services had to modify their practice and resources.^23-25^ These changes may have impacted HCC care practice as well, delaying diagnosis and/or treatment in some settings. This hypothesis is supported by data from 27 hospitals in 14 Asia-pacific countries, including Australia, indicating a 27% decrease in new HCC cases during early 2020 compared to 2019.^26^ About half of the hospitals reported delays in diagnosis of new HCC cases and/or initiating HCC treatment. Similarly, another international survey among 91 clinical centers from Europe, Americas, Asia and Africa indicated that 81% of liver cancer screening programs and 41% of diagnostic procedures were either deferred or cancelled in early 2020.^27^ This hypothesis, however, is not supported by our other findings which demonstrated that the proportion of HCC cases diagnosed at early stages and proportion of those eligible for curative HCC management increased or remained consistent after 2019. Second, the expansion of multidisciplinary teams and qualified clinical services to manage HCC in other hospitals may have resulted in decreasing number of patients referred to the referral hospitals involved in this study. Further studies, including continuous monitoring of the number of HCC cases in 2022 and 2023 can provide more information in this regard.

Our data demonstrated that following an initial steady increase, the proportion of HCV HCC cases almost halved during 2017-21 compared to pre-2017. The initial increase in HCV HCC cases is consistent with previous data, demonstrating the rising burden of HCV-related liver disease in Australia before 2016, including increased hospitalization and mortality among people with HCV infection.^28,29^ This pattern was mainly related to the “ageing cohort” effect in the population living with HCV, and the low uptake and efficacy of interferon-containing HCV treatment (previous standard of care for HCV), particularly in patients with advanced liver disease.^28^ Our findings of decreased HCV HCC cases since 2017 could be explained by universal access to HCV DAA treatment since March 2016,^14^ and consequently a rapid scale-up in DAA treatment uptake among people with advanced liver disease. It was estimated that about 70% of people with HCV-related cirrhosis received DAA treatment by the end of 2017.^14^ Our findings are consistent with another study demonstrating significant changes in the trend of decompensated cirrhosis and HCC diagnosis and mortality among people with HCV in NSW after access to DAA therapy.^11^

Despite decreased frequency of HCV HCC post-2016, our data identified that about 30% of HCC cases during 2017-21 were still HCV-related. A meta-analysis demonstrated that although HCV cure reduced the risk of HCC, it did not eliminate the risk, with the incidence of HCC after HCV cure estimated as 2.1 per 100 person-years among patients with cirrhosis and 0.5 per 100 person-years among patients with F3 fibrosis.^30^ These data, consistent with our findings, suggest that post-treatment HCC surveillance is required among people with HCV, including those with cirrhosis pre-cure. Our data also identified that 39% of people with HCV HCC had ARLD as a co-morbidity. Given the contribution of ARLD to liver disease progression, ^31^ alcohol use among people with HCV should be monitored given that it may compromise the benefits of DAA therapy.

Our findings identified that only 48% of patients with HBV HCC were receiving HBV treatment prior to HCC diagnosis. This finding is consistent with the sub-optimal treatment uptake among overall people living with HBV in Australia. In 2020, an estimated 11% of people with HBV in Australia were receiving HBV treatment,^32^ far lower than an estimated 18-20% of people living with chronic HBV eligible for treatment.^33^ Interventions to improve HBV treatment uptake required, which potentially impacts the rate of HCC as well.^12^

Our findings identified an increasing number and proportion of NAFLD/NASH HCC cases over time, although the trend was not statistically significant. Other studies also demonstrated an increasing burden of NASH HCC in several developed countries.^34-36^ In addition, a recent epidemiological study in South Australia described a 7% increase in NAFLD/NASH HCC cases from 2014-19, forecasting an increasing trend.^37^ These data, in line with our findings, suggest NAFLD/NASH as an emerging HCC risk factor in many countries, including Australia. Clinical and public health strategies are needed to properly respond to the increasing burden of metabolic syndrome, including NASH and NAFLD.

Our findings identified an increasing proportion of patients who were diagnosed at earlier HCC stages and receiving curative treatments. This trend could be explained by the increasing proportion of cases detected through HCC surveillance from 28% in 2008-09 to 42% in 2020-21. Although the trend was not statistically significant in our analysis, it should be interpreted conservatively. Our analysis was capable to assess a linear trend that is sensitive to fluctuations similar to that observed in the proportion of HCC cases detected through surveillance during 2017-20. Several studies demonstrated that HCC screening was associated with lower HCC burden and increased survival.^38-41^

Our findings identified that a higher proportion of patients with viral hepatitis-related HCC were diagnosed through surveillance compared to those with ARLD or NAFLD/NASH HCC. Our data also demonstrated higher proportion of HCC diagnosis at early stage among people receiving anti-viral treatment, regardless of response to treatment. Patients with viral hepatitis are more likely to be engaged with medical services due to the clinical care they may receive for their viral hepatitis. Given the increasing trend in NAFLD/NASH HCC cases observed in this study, strategies tailored for people with NAFLD/NASH or ARLD are needed to improve their engagement in clinical care, for both treatments of their underlying condition and HCC screening of eligible patients.

This study has several limitations. We used the data from three large public hospitals in NSW. Therefore, the study population may not be fully representative of patients from other settings. Data on ARLD and NAFLD/NASH were mainly based on documentation in clinical notes and subject to possible under-reporting, particularly in early years of the study period. Liver ablation and resection were considered curative HCC management methods although in some cases they may not have been curative. Apart from HCV HCC, our trend analyses were not able to accurately evaluate non-linear trends. Lastly, the impact of the COVID-19 pandemic was not fully explored in this study given the scope of the current analysis and unavailable required data.

In conclusion, this study demonstrated a decrease in the number of HCC cases after 2019, probably due to COVID-19-related restrictions. The number and proportion of patients with HCV HCC have been decreasing since DAA therapy became widely available, demonstrating the role of HCV elimination in decreasing HCC risk at population-level. The proportion of patients presenting with early-stage tumors has been increasing. Current healthcare policies should continue to promote HCV treatment and HCC surveillance following HCV cure among eligible patients. Moreover, other strategies should be developed for the prevention and management of emerging HCC risk factors including NAFLD/NASH.

## Data Availability

The data that support the findings of this study are available on request from the corresponding author. The data are not publicly available due to privacy or ethical restrictions.

## Acknowledgements

The Kirby Institute is funded by the Australian Government Department of Health and Ageing. The views expressed in this publication are those of the authors and do not necessarily represent the position of the Australian Government.

## Supplementary materials

**Figure 1:**
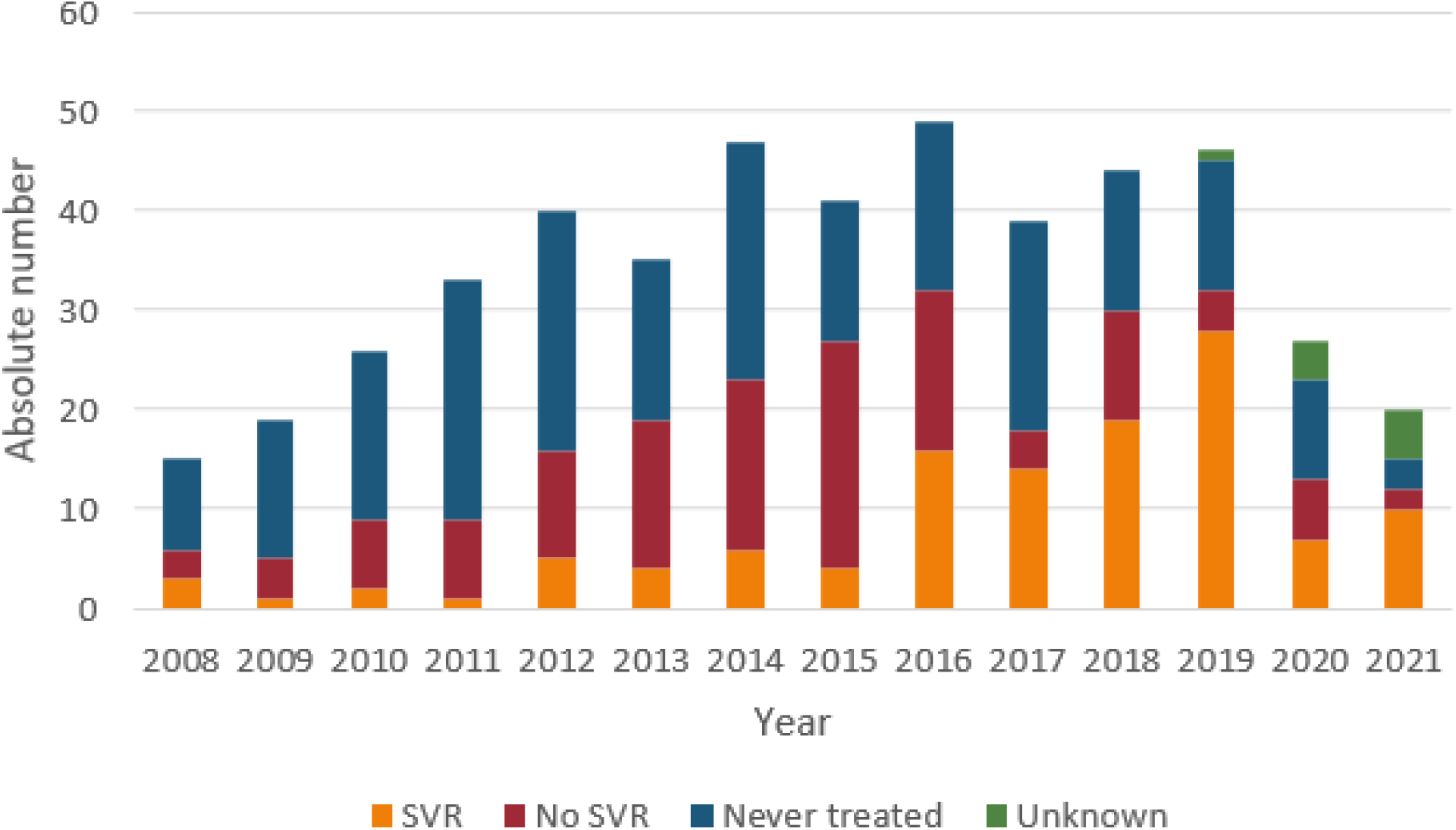
Classification of HCV treatment in HCV HCC patients, by year of HCC diagnosis. (SVR= sustained virologic response, HCV= hepatitis C, HCC= hepatocellular carcinoma)

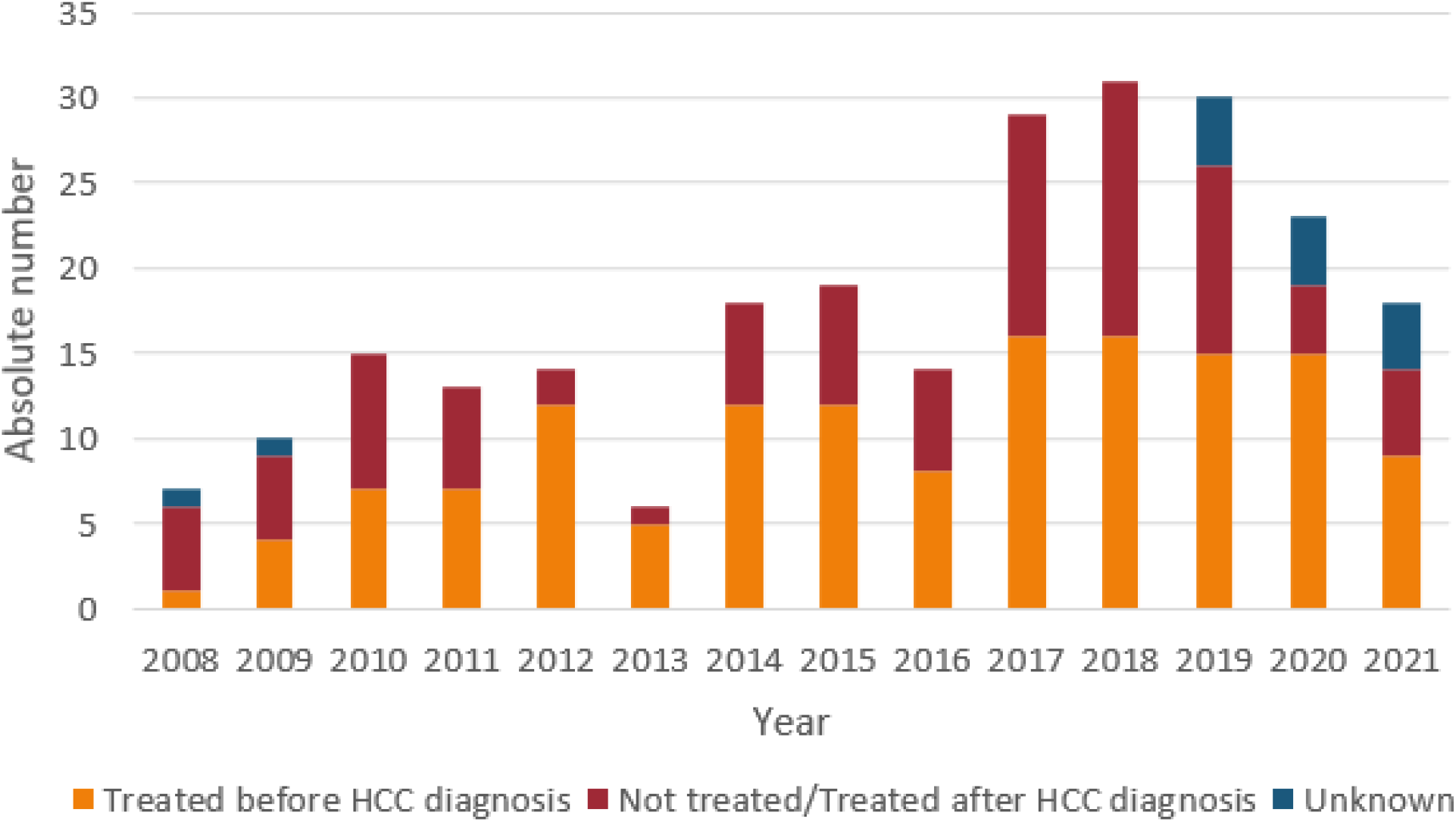

